# BrainAGE Estimation: Influence of Field Strength, Voxel Size, Race, and Ethnicity

**DOI:** 10.1101/2023.12.05.23299222

**Authors:** Desarae A. Dempsey, Rachael Deardorff, Yu-Chien Wu, Meichen Yu, Liana G. Apostolova, Jared Brosch, David G. Clark, Martin R. Farlow, Sujuan Gao, Sophia Wang, Andrew J. Saykin, Shannon L. Risacher, the Alzheimer’s Disease Neuroimaging Initiative

**Author notes:** <u>Corresponding Author</u> Shannon L. Risacher, PhD, Associate Professor of Radiology & Imaging Sciences, Center for Neuroimaging, Department of Radiology and Imaging Sciences, Indiana Alzheimer’s Disease Research Center, 355 West 16^th^ Street, Suite 4100, Indianapolis, IN 46202 USA. Data used in preparation of this article were obtained from the Alzheimer’s Disease Neuroimaging Initiative (ADNI) database (adni.loni.usc.edu). As such, the investigators within the ADNI contributed to the design and implementation of ADNI and/or provided data but did not participate in analysis or writing of this report. A complete listing of ADNI investigators can be found at: http://adni.loni.usc.edu/wp-content/uploads/how_to_apply/ADNI_Acknowledgement_List.pdf.

## Abstract

The BrainAGE method is used to estimate biological brain age using structural neuroimaging. However, the stability of the model across different scan parameters and races/ethnicities has not been thoroughly investigated. Estimated brain age was compared within- and across-MRI field strength and across voxel sizes. Estimated brain age gap (BAG) was compared across demographically matched groups of different self-reported races and ethnicities in ADNI and IMAS cohorts. Longitudinal ComBat was used to correct for potential scanner effects. The brain age method was stable within field strength, but less stable across different field strengths. The method was stable across voxel sizes. There was a significant difference in BAG between races, but not ethnicities. Correction procedures are suggested to eliminate variation across scanner field strength while maintaining accurate brain age estimation. Further studies are warranted to determine the factors contributing to racial differences in BAG.

## 1. Introduction

Brain age estimation techniques have become popular over the last decade as potential biomarkers of individualized brain health and age-related disease severity or prediction [1-5]. The brain age algorithm uses a machine learning model, typically in the form of a regression, trained on structural magnetic resonance imaging (MRI) scans from a large group of healthy individuals from a wide age range to generate a typical pattern of brain aging [1]. The algorithm can then be applied on independent datasets to predict the brain age of individuals, and the deviation from their chronological age can be calculated, which is referred to as the brain age gap (BAG) [1].

Brain aging measures are specifically relevant in patients with Alzheimer’s disease and related dementias (ADRD) due to accelerated brain atrophy in these disorders and the relationship between brain atrophy and cognitive decline [6-11]. The brain age technique has been previously used to measure brain aging in cognitively normal (CN), mild cognitive impairment (MCI), and ADRD [7, 8, 12-19]. Results show that brain age techniques are a potential biomarker for detecting neuroanatomical brain changes that could facilitate early detection before the presentation of clinical AD symptoms [5, 8].

Previous studies have uncovered some biases and limitations in the brain age method. Studies have shown that the brain age framework inherently underestimates the age of older individuals and overestimates the age of younger individuals, compared to their chronological age; corrected procedures are suggested [3, 20-27]. Studies have also provided evidence that the brain age model is stable within- and across-scanner field strength, reported using intraclass correlation coefficient (ICC) values as a measure of test-retest repeatability [2, 3, 12]. However, a linear offset for field strength has been observed [2, 12]. To date, there are no studies comparing stability across different scan voxel sizes, which has been noted as a limitation [3]. Another noted limitation of the brain age model is that majority of studies are conducted in primarily or exclusively non-Hispanic White race participants [1], although a few brain age studies have been conducted in East Asian populations [13, 21, 28]. To our knowledge, there are no studies comparing racial or ethnic groups using the brain age algorithm despite previous studies suggesting that evidence of racial, ethnic, or cultural differences are represented in brain structure [1, 29-36].

The overall aim of this study was to define ways to harmonize the brain age algorithm output in large datasets containing scans with different parameters and individuals from different racial and ethnic groups. Thus, we had two main goals. The first was to determine whether the brain age estimations are stable across different scanner/scan parameters, namely field strength and voxel size. To assess field strength, we used participants from the Alzheimer’s Disease Neuroimaging Initiative (ADNI) with multiple structural MRI scans in both a 1.5T scanner and a 3T scanner and performed a repeatability analysis within- and across-scanner field strength. Then, we assessed structural MRI scans at three different inherent voxel sizes from participants in the Indiana Memory and Aging Study (IMAS) and performed repeatability analysis across the different voxel sizes. To attempt data harmonization across scanner parameters, we investigated the use of ComBat. ComBat harmonization is a tool that was first created to eliminate batch effects for gene expression microarray data [37]. ComBat has been translated into use for both cross-sectional and longitudinal multi-modal neuroimaging data [38-41]. ComBat is an empirical Bayes method that is useful for removing unwanted technical variability across scanners and sites, while preserving biological variability, and is robust for small sample sizes [37, 38]. Our second goal was to investigate differences in BAG between individuals from different racial or ethnic groups. To accomplish this goal, we assessed differences in BAG between demographically matched African American (AA) individuals and non-Hispanic White (NHW) individuals from IMAS, as well as demographically matched AA and NHW, Asian and NHW, and Hispanic White (HW) and NHW individuals from ADNI.

## 2. Methods

The data presented within this article were from participants from both ADNI (http://adni.loni.usc.edu) and IMAS. These studies are discussed in detail in the Supplemental text. Details of the sub-samples from each cohort that were used for each parameter investigation are described below. All MRI scans used were 3D T1-weighted magnetization-prepared rapid gradient-echo (MPRAGE) scans. Individuals and single scans were excluded for failed BrainAGE processing after visual quality control. Outliers of 3 standard deviations above or below the mean BAG were also excluded.

### 2.1. Consent Statement

Informed consent was obtained according to the Declaration of Helsinki.

### 2.2. ADNI – Field Strength Analysis

ADNI is a multi-site longitudinal study designed to improve and standardize neuroimaging biomarkers for studying AD. For more details, see the Supplementary text. For the field strength analyses, ADNI participants from the ADNI-1 phase who had undergone at least two 1.5T MPRAGE scans in the same session and at least two 3T MPRAGE scans in a separate single session were included. The duplicate scans in different scanner types were conducted on average a few weeks apart. AD and MCI were diagnosed using standard criteria [42, 43]. CN participants had no significant memory concerns or performance deficits on cognitive testing. 25 individuals from each diagnostic group (CN, MCI, AD) were randomly selected and included in the analyses, for a total of 75 participants. All participants were NHW, due to uncertainty of brain age model accuracy on other racial and ethnic groups. 3D T1-weighted MPRAGEs were acquired on various scanners collected at over 50 sites. All scans followed the ADNI-1 MPRAGE sequence (see http://adni.loni.usc.edu). Pre-processed MRI scans were downloaded from ADNI and processed with using the brain age algorithm as described below.

### 2.3. IMAS – Voxel Size Analysis

The Indiana Memory and Aging Study (IMAS) is a longitudinal observational study of older adults at risk for and with clinical AD. For more details on IMAS and diagnostic criteria, see the Supplemental text and references [42-47]. Included individuals had at least two MPRAGE scans of different inherent voxel sizes collected within the same scan session. Participants had multiple 3D T1-weighted MPRAGEs acquired on a Siemens Prisma 3T scanner at 3 varying inherent voxel sizes: 1) 0.8mm^3^, 2) 1mm^3^, 3) 1.1mm x 1.1mm x 1.2mm (termed “ADNI2 sequence”). Specifically, 48 NHW participants (19 CN, 16 SCD, 10 MCI, 3 AD) were included in the analyses, including 47 participants with one 0.8mm^3^ scan and one ADNI2 sequence scan, and 16 participants with all three scans (one 0.8mm^3^, one 1mm^3^, one ADNI2 sequence). Notably, we also tested whether preregistering the scans to Montreal Neurologic Institute (MNI) space affected the brain age results compared to native space scans, which it did not (ICC=0.999 (p<0.001); R^2^ =0.999 (p<0.001); *data not shown*).

### 2.4. Race/Ethnicity Comparisons (ADNI and IMAS)

In IMAS, 71 AA individuals were each matched as closely as possible on sex, age, education, diagnostic group, and handedness to a NHW individual. All IMAS scans were performed on the same Siemens Prisma 3T scanner and processed using the BrainAGE algorithm described below. In ADNI, 147 AA, 76 HW, and 47 Asian participants were each matched as closely as possible on sex, age, education, diagnostic group, magnet field strength, and handedness to a NHW participant from the same phase of ADNI (ADNI1, ADNIGO/2, ADNI3). Pre-processed 3D MPRAGE scans were downloaded from ADNI (http://adni.loni.usc.edu) and processed using the BrainAGE algorithm as described below.

### 2.5. Brain Age Framework

Predicted raw brain age estimations were generated using brainageR, (version 1.0, https://github.com/james-cole/brainageR/releases/tag/1.0), which is performed in R and uses SPM12 and KernLab [3, 14, 48, 49]. A machine-learning Gaussian Process Regression (GPR) was used, which was based on a training dataset of structural MRI scans of 2001 healthy individuals (male/female: 1016/985, mean age: 36.95 (18.12), age range: 18-90 years), to capture the typical brain aging trajectory over time [3, 14]. This GPR was created from multiple public access data sets using different collection sites, various scanners including both 1.5T and 3T, and various voxel sizes [3, 14]. Each participant’s T1-weighted MRI scan from our data set was then registered to standard space, segmented, and compared to this GPR. Then, the code provided an estimated brain age for each participant based on his/her/their brain structure. The BAG is then calculated by subtracting an individual’s chronological age from his/her/their brain age estimation to measure the residual or distance from the GPR, which is a measure of the individual’s deviation from the typical brain aging trajectory. A positive BAG indicates accelerated aging, while a negative BAG indicates decelerated aging compared to the typical brain aging trajectory [2]. For example, an individual that is 67 years old chronologically, but has a brain resembles that of a typical 70-year-old, will have a raw brain age estimation of 70, and a BAG of +3 years, indicating accelerated aging [2].

### 2.6. ComBat Harmonization Method

We used longitudinal ComBat to remove the technical variability seen across scanner field strengths, while maintaining the biological variability in the brain age estimation. Longitudinal ComBat was designed to correct for additive and multiplicative scanner effects while accounting for the within-subject correlation inherent to longitudinal data [38]. Longitudinal ComBat was performed on the brain age values from the repeat 1.5T scans and the repeat 3T scans. The R code for longitudinal ComBat can be found at https://github.com/jcbeer/longCombat [38].

### 2.7. Statistical Analyses

Statistical analyses were performed using R studio / R version 4.2.1. The raw brain age estimations and BAG scores are reported as mean ± standard deviation in years.

For field strength and voxel-size analyses, intraclass correlation coefficients (ICC: two-way random-effects, absolute agreement, single rater/measurements) and regressions reporting R-squared (R^2^) values were calculated to assess the repeatability of the brain age output within- and across-scans of different field strengths and across scans of different voxel sizes [3, 45]. In addition, paired t-tests were calculated to determine whether mean brain age estimates were significantly different between scans with different scanner parameters. For the within field strength analyses, a p-value < 0.05 is considered significant. For the across field strength analyses, if Bonferroni correction is applied, a p-value < 0.0125 is considered significant. All p-values far surpassed this cut off for multiple comparison testing, so we report the raw p-values as they are all significant after correction. If Bonferroni correction is applied for the voxel size analyses, a p-value < 0.017 is considered significant.

For the race/ethnicity analyses, independent group t-tests (two-tailed) were used to compare the mean BAG between self-reported racial or ethnic groups. To check appropriate demographic matching, demographic variables were compared between racial groups using a Chi-Square test for nominal variables (diagnostic group, sex, handedness) or an independent t-test (for continuous variables (years of education, scan age)). For the race/ethnicity analyses, no correction is necessary for the independent analysis comparison of AAs and NHWs in the IMAS cohort, and a p-value < 0.05 is considered significant. For the ADNI cohort, each comparison may be considered independent, but since there is potential overlap between the NHW groups, we acknowledge that if Bonferroni correction is applied, a p-value < 0.017 is considered significant.

## 3. Results

### 3.1. Comparisons of Estimated Brain Age within Field Strength

Repeatability scores were consistently high between scans within the same scanner parameters. Raw brain age estimations from two 1.5T scans showed good consistency with ICC=0.992 and R^2^=0.984 (both p<0.001; Figure 1A, Table 1). Similarly, raw brain age estimations from two 3T scans were consistent with an ICC of 0.987 and R^2^ =0.975 (both p<0.001; Figure 1B, Table 1). The 1.5T scans gave mean brain age estimates of 74.62 ± 9.69 years and 74.74 ± 9.94 years, while the 3T scans gave mean brain age estimates of 70.49 ± 9.87 years and 70.02 ± 9.45 (Supplemental Table 1). No significant difference within field strength was observed for either the 1.5T or 3T scans (p>0.05; Table 1).

**Table 1.** Comparisons Within and Across Scanner Field Strength in ADNI ICC, R^2^ values, and paired T-test results for the brain age results are shown for each comparison. There was no significant difference within scanner field strength, but there was a significant difference between every pair across scanner field strength. After ComBat and after linear transformation, there was no longer a significant difference between the brain age values across field strengths. ICC = intraclass correlation coefficient.

| Comparison (x,y) |  | Linear Equation | ICC (p-value) | R<sup>2</sup> (p-value) | T-statistic (p-value) |
| --- | --- | --- | --- | --- | --- |
| Within Field Strength |  |  |  |  |  |
| 1.5T Scan #1 | 1.5T Scan #2 | $y = 1.01x - 0.35$ | 0.992 (p<0.001) | 0.984 (p<0.001) | -0.264 (p=0.793) |
| 3T Scan #1 | 3T Scan #2 | $y = 0.99x + 0.73$ | 0.987 (p<0.001) | 0.975 (p<0.001) | 0.667 (p=0.507) |
| Across Field Strength |  |  |  |  |  |
| 1.5T Scan #1 | 3T Scan #1 | $y = 0.96x - 1.11$ | 0.865 (p<0.01) | 0.886 (p<0.001) | 10.680 (p<0.001) |
| 1.5T Scan #1 | 3T Scan #2 | $y = 0.95x - 0.49$ | 0.840 (p<0.01) | 0.859 (p<0.001) | 10.194 (p<0.001) |
| 1.5T Scan #2 | 3T Scan #1 | $y = 0.94x + 0.05$ | 0.870 (p<0.01) | 0.888 (p<0.001) | 10.385 (p<0.001) |
| 1.5T Scan #2 | 3T Scan #2 | $y = 0.94x + 0.48$ | 0.848 (p<0.01) | 0.865 (p<0.001) | 10.004 (p<0.001) |
| All 1.5T Scans | All 3T Scans | $y = 0.95x - 0.35$ | 0.856 (p<0.01) | 0.876 (p<0.001) | 14.665 (p<0.001) |
| Across Field Strength - After ComBat |  |  |  |  |  |
| 1.5T Scan #1 | 3T Scan #1 | $y = 0.94x + 4.10$ | 0.926 (p<0.001) | 0.856 (p<0.001) | -0.078 (p=0.938) |
| 1.5T Scan #1 | 3T Scan #2 | $y = 0.93x + 4.91$ | 0.908 (p<0.001) | 0.822 (p<0.001) | 0.171 (p=0.865) |
| 1.5T Scan #2 | 3T Scan #1 | $y = 0.93x + 5.31$ | 0.928 (p<0.001) | 0.859 (p<0.001) | -0.145 (p=0.885) |
| 1.5T Scan #2 | 3T Scan #2 | $y = 0.92x + 5.89$ | 0.912 (p<0.001) | 0.831 (p<0.001) | 0.046 (p=0.963) |
| Across Field Strength – After Linear Transformation |  |  |  |  |  |
| 1.5T Scan #1 | 3T Scan #1 | $y = 1.00x - 0.80$ | 0.940 (p<0.001) | 0.866 (p<0.001) | 0.138 (p=0.891) |
| 1.5T Scan #1 | 3T Scan #2 | $y = 1.00x - 0.15$ | 0.925 (p<0.001) | 0.856 (p<0.001) | 0.438 (p=0.663) |
| 1.5T Scan #2 | 3T Scan #1 | $y = 0.99x + 0.42$ | 0.942 (p<0.001) | 0.888 (p<0.001) | 0.030 (p=0.977) |
| 1.5T Scan #2 | 3T Scan #2 | $y = 0.99x + 0.87$ | 0.929 (p<0.001) | 0.856 (p<0.001) | 0.280 (p=0.780) |
| All 1.5T Scans | All 3T Scans | $y = 1.00x - 0.01$ | 0.934 (p<0.001) | 0.876 (p<0.001) | 0.298 (p=0.766) |

**Figure 1.**
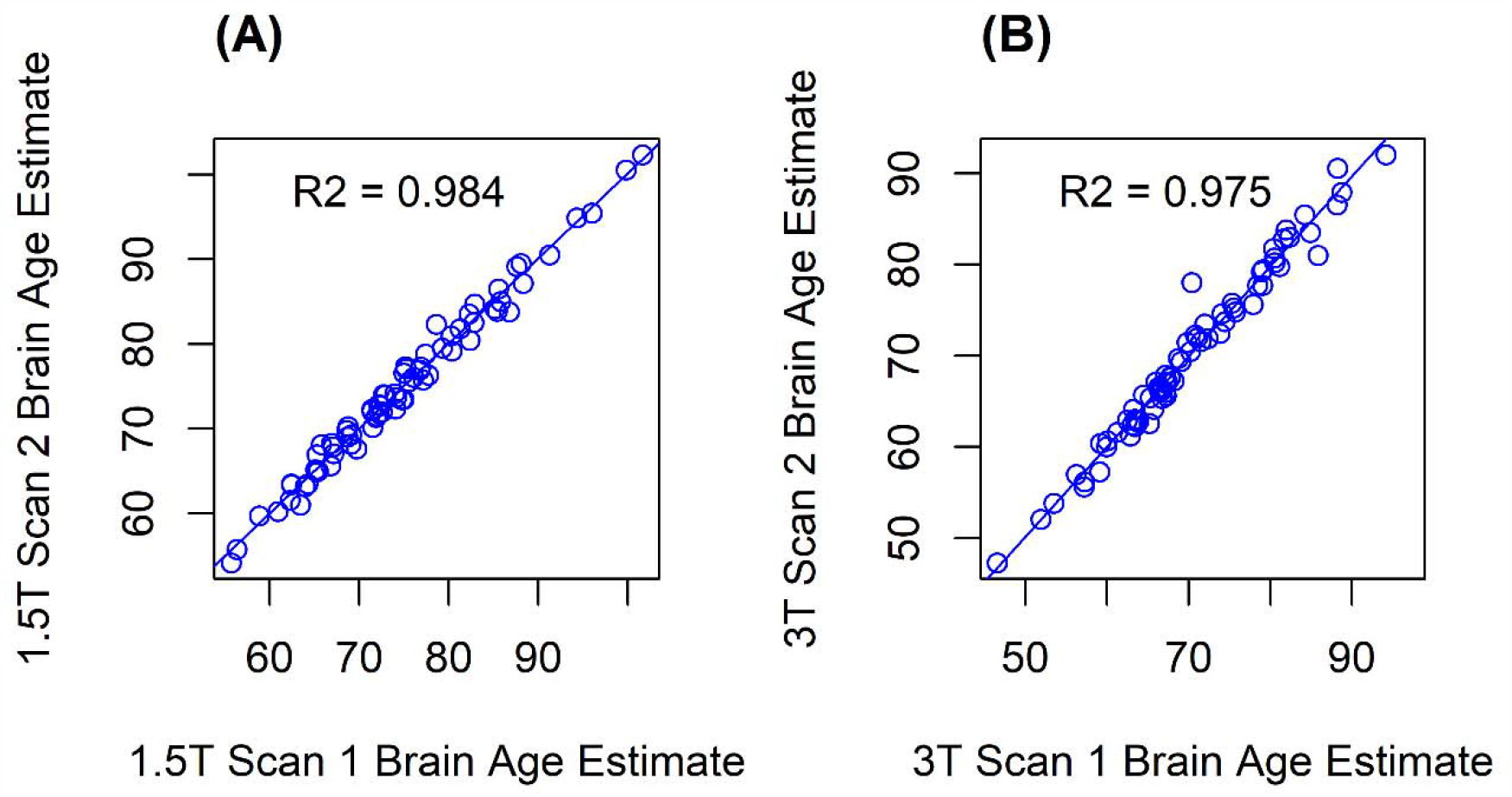
Brain Age Estimation Within Field Strength Scatterplots showing the association of raw brain age estimation values **(A)** between scans 1 and 2 within the 1.5T field strength showed an ICC=0.992 (p< 0.001) and R^2^=0.984 (p<0.001), with the linear equation: y=1.01x-0.35. **(B)** The association between scan 1 and 2 within the 3T field strength shows an ICC=0.987 (p<0.001) and R^2^=0.975 (p<0.001), with the linear equation: y=0.99x+0.73. ICC = intraclass correlation coefficient

### 3.2 Comparisons of Brain Age Estimates across Field Strength

When comparing across scans from different MRI field strength (1.5T vs 3T), the brain age estimation tool was less consistent than within field strength. There were two 1.5T scans and two 3T scans for each participant, thus there were four possible comparisons: 1.5T scan 1 vs. 3T scan 1, 1.5T scan 1 vs 3T scan 2, 1.5T scan 2 vs 3T scan 1, and 1.5T scan 2 vs 3T scan 2. The ICCs for these comparisons ranged from 0.840 – 0.870 (all p<0.01, Table 1), while R^2^ values for these comparisons ranged from 0.859 – 0.888 (all p<0.001; Figure 2, Table 1). Linear equations and statistical results describing the brain age estimate relationship between 1.5T and 3T scans are shown in Table 1. Mean brain age estimates for each scan type are shown in Supplemental Table S1 and ranged from 70.02 ± 9.45 for 3T scan 2 to 74.62 ± 9.69 for 1.5T scan 1. Brain age estimates were significantly different for all four comparisons (p<0.001; Table 2). The 3T scanner consistently estimated a lower brain age than the 1.5T scanner, with an offset of 4.43 years on average.

**Table 2.** Comparisons Across Scanner Voxel Size in IMAS.

| Comparison (x,y) |  | Linear Equation | ICC (p-value) | R<sup>2</sup> (p-value) | T-statistic (p-value) |
| --- | --- | --- | --- | --- | --- |

| Across Voxel Size |  |  |  |  |  |
| --- | --- | --- | --- | --- | --- |
| 0.8mm<sup>3</sup> Scan | 1mm<sup>3</sup> Scan | $y = 0.83x + 11.94$ | 0.968 (p<0.001) | 0.965 (p<0.001) | 1.011 (p=0.328) |
| 0.8mm<sup>3</sup> Scan | ADNI2 Scan | $y = 0.90x + 7.66$ | 0.929 (p<0.001) | 0.865 (p<0.001) | -1.166 (p=0.250) |
| 1mm<sup>3</sup> Scan | ADNI2 Scan | $y = 1.10x - 5.78$ | 0.965 (p<0.001) | 0.957 (p<0.001) | -2.249 (p=0.040) |

ICC, $R^2$ values, and paired T-test results for the brain age results are shown for each comparison.
There was a difference between the brain age results from the 1mm<sup>3</sup> scan and the ADNI2 scan.
There was no significant difference between the 0.8mm<sup>3</sup> scan and the 1mm<sup>3</sup> scan or the 0.8mm<sup>3</sup> scan and the ADNI2 scan. ICC = intraclass correlation coefficient

**Figure 2.**
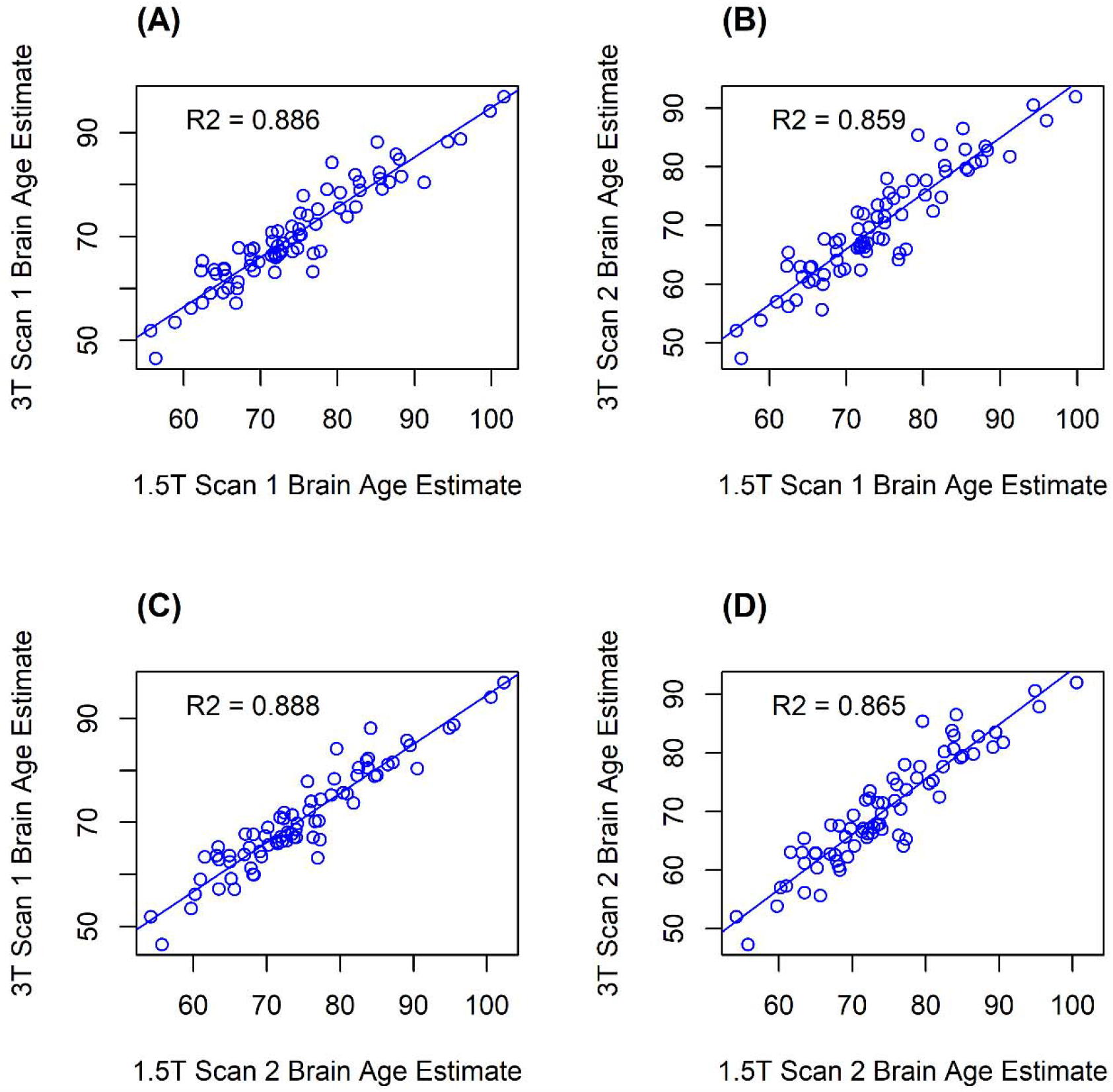
Brain Age Estimation Across Field Strength Scatterplots showing the correlation of raw brain age estimation values **(A)** between 1.5T scan 1 and 3T scan 1, resulting in repeatability values of: ICC=0.865 (p<0.01) and R^2^=0.886 (p<0.001), with the linear equation: y=0.96x-1.11 **(B)** between 1.5T scan 1 and 3T scan 2, resulting in repeatability values of: ICC=0.840 (p<0.01) and R^2^=0.859 (p<0.001), with the linear equation: y=0.95x-0.49 **(C)** between 1.5T scan 2 and 3T scan 1, resulting in repeatability values of: ICC=0.870 (p<0.01) and R^2^=0.888 (p<0.001), with the linear equation: y=0.94x+0.05 **(D)** between 1.5T scan 2 and 3T scan 2, resulting in repeatability values of: ICC=0.848 (p<0.01) and R^2^=0.865 (p<0.001), with the linear equation: y=0.94x+0.48. ICC = intraclass correlation coefficient

### 3.3. Comparisons of Brain Age Estimates across Voxel Size

Similar to the field strength analyses above, three different comparisons were calculated to assess differences in raw brain age estimation by voxel size: 0.8mm^3^ scan vs. 1mm^3^ scan, 0.8mm^3^ scan vs. ADNI2 scan (1.1x1.1x1.2mm), and 1mm^3^ scan vs. ADNI2 scan. The ICC value for the comparison of the 0.8mm^3^ scan to the 1mm^3^ scan was 0.968 and the R^2^ value was 0.965 (all p<0.001; Figure 3A, Table 2). The ICC value for the comparison of the 0.8mm^3^ scan to the ADNI2 scan was 0.929 and the R^2^ value was 0.865 (all p<0.001; Figure 3B, Table 2). Finally, the comparison between the 1mm^3^ scan and the ADNI2 scan had an ICC of 0.965 and R^2^ of 0.957 (both p<0.001; Figure 3C, Table 2), The mean brain age estimation for 0.8mm^3^ scan was 70.69 ± 11.05 years, for the 1mm^3^ scan was 71.64 ± 8.99 years, and for the ADNI2 scan was 71.13 ± 10.45 years (Supplementary Table S2). No significant differences were observed between the mean brain age estimates between the 0.8mm^3^ and the 1mm^3^ scans, or the 0.8mm^3^ and the ADNI2 scans (p>0.05; Table 2). However, a trend for a difference between 1mm^3^ and ADNI2 scans was observed but did not meet corrected significance (p=0.04; Table 2).

**Figure 3.**
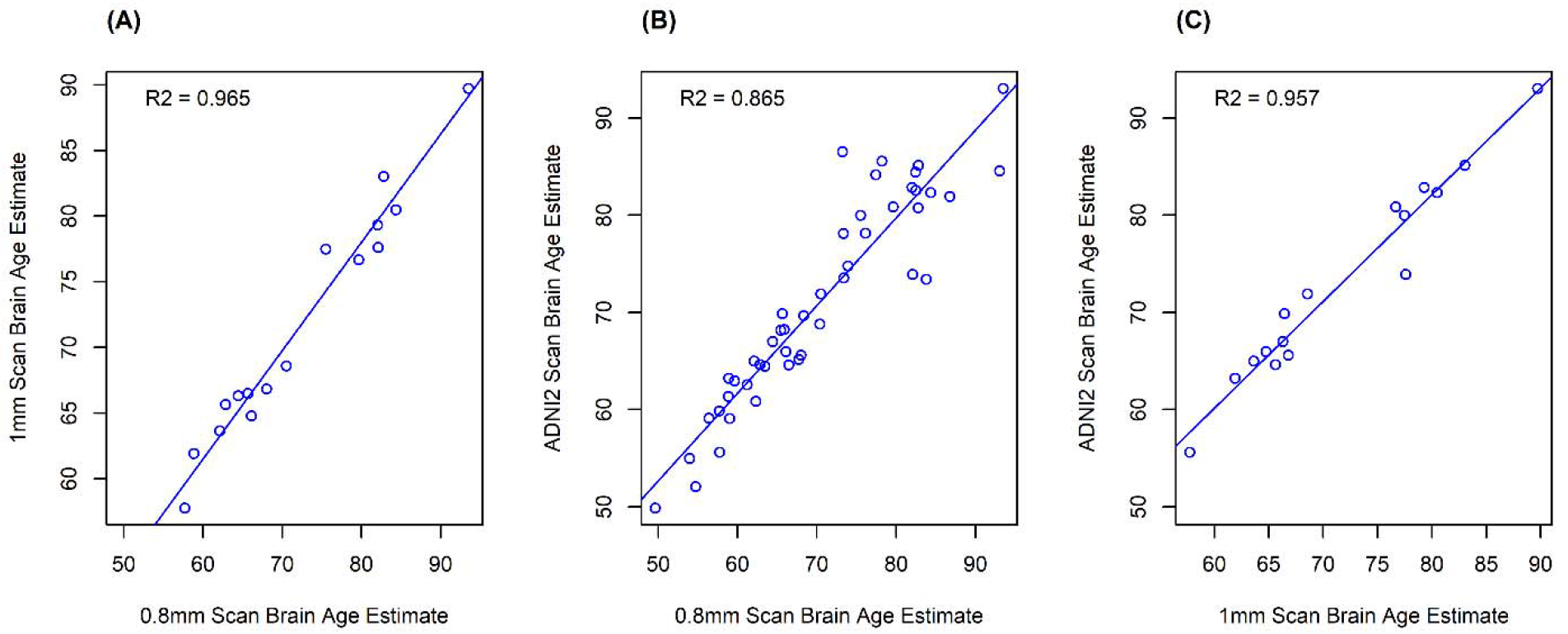
Brain Age Estimation Across Voxel Size Scatterplots showing the correlation of the raw brain age estimation values **(A)** between the 0.8mm^3^ scan and the 1mm^3^ scan, resulting in repeatability values of: ICC=0.968 (p<0.001) and R^2^=0.965 (p<0.001), with the linear equation: y=0.83x+11.94 **(B)** between the 0.8mm^3^ scan and the ADNI2 scan, resulting in repeatability values of: ICC=0.929 (p<0.001) and R^2^=0.865 (p<0.001), with the linear equation: y=0.90x+7.66 **(C)** between the 1mm^3^ scan compared to the ADNI2 sequence (1.1x1.1x1.2mm), resulting in repeatability values of: ICC=0.965 (p<0.001) and R^2^=0.957 (p<0.001), with the linear equation: y=1.10x-5.78. ICC = intraclass correlation coefficient

### 3.4. ComBat Harmonization Results

Additive effects but no multiplicative effects were found, so longitudinal ComBat was applied for correction. After longitudinal ComBat, there was no longer an additive effect nor significant differences in brain age estimations across field strengths (p>0.05, Table 1). Using the ComBat batch-corrected values, the ICC values increased, while the R^2^ values slightly decreased. The details are shown in Table 1.

### 3.5 Linear Transformation Results

A linear transformation equation was used to remove differences across scanner field strengths. This linear equation was created by pooling all brain age values from the 1.5T scans and regressing these against all the 3T scans. The regression equation produced was y = 0.95x –0.35. We then applied this equation (adjusted 3T estimate = (raw 3T estimate + 0.35) / 0.95) to all 3T scan brain age values to correct for the scanner difference. After application of this linear transformation, no significant differences across scanner field strength were observed. Further, the ICC values increased, while the R^2^ values remained the same relative to the original analyses. The details are shown in Table 1.

### 3.6. Comparisons of Brain Age Gap between Demographically Matched Racial and Ethnic Groups

To test BAG between racial and ethnic groups, we first calculated each participant’s BAG (brain age estimate - chronological age). We then compared the mean BAG of each racial or ethnic group (AA, HW, or Asian) to a sample of NHWs matched on diagnostic group, sex, age, education, and handedness, as well as ADNI phase for the ADNI analyses. No significant differences in any demographic variables were detected between racial or ethnic groups, which confirmed appropriate demographic matching (Supplementary Tables S3-S6).

In IMAS, mean BAG was significantly different between NHWs (-3.87 ± 6.79 years) and AA (- 6.49 ± 8.50 years) (t=2.03, p=0.04; Figure 4A, Supplemental Table S3). A similar effect was seen in the ADNI cohort, with a significant difference in mean BAG between the NHW group (- 3.87 ± 8.84 years) and the AA group (-6.79 ± 8.01 years) observed (t=2.96, p=0.003; Figure 4B, Supplemental Table S4). However, BAG was not significantly different between the NHW group (-2.76 ± 8.04 years) and the HW group (-3.75 ± 7.64 years) (t=0.77, p=0.441; Figure 4C, Supplemental Table S5). Finally, a significant difference in BAG between the NHW group (- 1.56 ± 9.64 years) and the Asian group (-5.59 ± 7.80 years) was observed for the raw p-value (t=2.22, p=0.029; Figure 4D, Supplemental Table S6), but not after correction.

**Figure 4.**
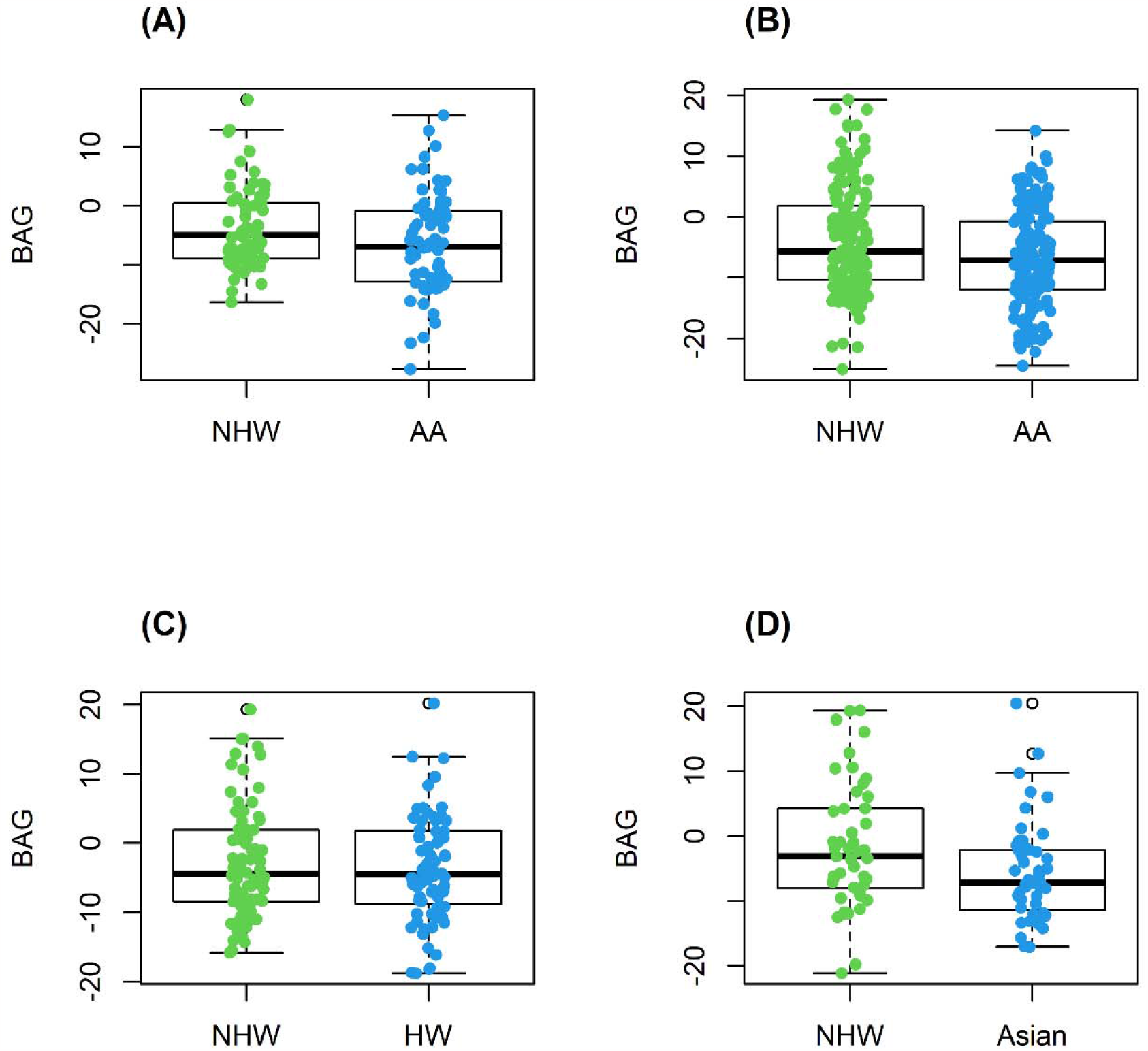
Brain Age Gap (BAG) Between Race and Ethnicity Groups Box plots comparing BAG between racial and ethnic groups matched on diagnostic group, sex, age, and handedness. **(A)** In the IMAS cohort, a significant difference in BAG between the Non-Hispanic White (NHW) group and the African American (AA) group was observed (t=2.03, p=0.04). **(B)** In the ADNI cohort, there was a significant difference in BAG between NHW and AAs (t=2.96, p=0.003) but **(C)** no difference in BAG between NHWs and Hispanic Whites (HWs) (t=0.77, p=0.441). **(D)** Finally, in ADNI, a significant difference in BAG was observed between NHWs and Asians (t=2.22, p=0.03), which did not remain after correction.

## 4. Discussion

The BrainAGE method was consistent across multiple scans of the same individual in the same scanner, but relatively less stable across scans of the same individual in different scanners with different field strengths. Brain age estimates from scans acquired in a 3T scanner estimated individuals to be ∼4.43 years younger than the estimates from scans acquired on a 1.5T scanner. This finding aligns with previous studies, where stability was lower across field strengths, and it was suggested that researchers correct for the field strength effect by shifting the BAG scores to a zero group mean [3, 12]. Longitudinal ComBat performed relatively well for the correction of scanner field strength. Using ComBat, the need for covarying for scanner is no longer necessary. In addition to the ComBat method, we propose a linear transformation (adjusted 3T estimate = (raw 3T estimate + 0.35) / 0.95), as this equation may be a better fit for correcting for scanner field strength, as it produced higher repeatability values in this study. In addition to supporting the differences by field strength, our results show for the first time that the brain age method was stable across scans of different voxel sizes. However, our group of participants with 1mm^3^ scans was rather limited (n=16), and thus, this analysis should be repeated in a larger sample, as well as with scans from other inherent voxel sizes.

To our knowledge, this paper is also the first to investigate differences in the brain age method by race or ethnicity. Our results show significant differences in BAG between racial groups (NHWs vs. AA, NHWs vs. Asians), but not between ethnic groups (NHWs vs. Hispanic Whites). However, it is noted that the Asian group did not have a significantly different BAG from NHWs if multiple comparison correction is applied. Both the AA group and the Asian group had lower BAG scores than respective demographically matched NHWs, suggesting decelerated aging in these groups. Previous studies have suggested that there are biological brain differences between East Asian and White, as well as AA and White CN individuals [29-33]. Studies have also suggested differences in presentation of AD and MCI with regard to both cognition and affected brain structures across NHWs, HWs, and AAs [34-36]. However, in the present study, we cannot determine whether the observed differences are biological or a bias from the machine learning model as most of the training set were NHWs. Additional studies should be conducted using larger multi-racial and multi-ethnic cohorts to clearly understand the different results across races from brain age techniques. Further, studies using the brain age technique in samples with mixed races should consider the impact of these factors when performing brain age analyses. Finally, we propose that separate machine learning models should be created and trained on subset of racial groups to ensure accuracy of brain age estimation. ADNI4 will be a prospective cohort that can be used to investigate racial differences further, as it will include a higher percentage of participants from underrepresented backgrounds.

One limitation of our study is the relatively small subgroup sample sizes, especially for the 1mm^3^ voxel size group (n=16). In addition, all cohorts have a high mean education level, which has been associated with lower brain age results [50]. We did not account for physical activity, which has previously been associated with lower brain age results as well [50]. Finally, while the basis for these findings is still unclear, the observed differences in BAG by race raises several important questions for future research on the neuroimaging methodologies in racial and ethnic groups and highlights the important need for improved recruitment of underrepresented minorities in AD research to better understand aging and age-related diseases in all individuals.

In conclusion, the brain age method was stable within field strength and across inherent voxel sizes. Brain age estimations are dependent on field strength, so we suggest using an offset equation or ComBat to harmonize data in studies with scans from scanners of different field strengths. Lastly, we found differences in BAG scores between self-reported races but are uncertain whether these represent biological differences or bias inherent in the algorithm. Further studies should explore this important area in larger multi-racial samples recruited from population-based studies.

## Data Availability

All data produced in the present study are available upon reasonable request.

## Acknowledgements

The authors thank Steve Brown, Bobi Eastman, Tatiana M. Foroud, Yolanda Graham-Dotson, Tyler Gannon, Savannah Hottle, Gary D. Hutchins, Brenna McDonald, Fredrick Unverzagt, Aaron Vosmeier, and Donna Wert for their contributions to this work.

## Collaborators

*Data used in preparation of this article were obtained from the Alzheimer’s Disease Neuroimaging Initiative (ADNI) database (adni.loni.usc.edu). As such, the investigators within the ADNI contributed to the design and implementation of ADNI and/or provided data but did not participate in analysis or writing of this report. A complete listing of ADNI investigators can be found at: http://adni.loni.usc.edu/wp-content/uploads/how_to_apply/ADNI_Acknowledgement_List.pdf

