## Supplemental for "BrainAGE Estimation: Influence of Field Strength, Voxel Size, Race, and Ethnicity"

**Supplemental Text**

**ADNI Methods**

ADNI was launched in 2003 by the National Institute on Aging (NIA), the National Institute of Biomedical Imaging and Bioengineering (NIBIB), the Food and Drug Administration (FDA), private pharmaceutical companies and non-profit organizations, as a $60 million, 5-year public-private partnership. The primary goal of ADNI has been to test whether serial magnetic resonance imaging (MRI), positron emission tomography (PET), other biological markers, and clinical and neuropsychological assessment can be combined to measure the progression of mild cognitive impairment (MCI) and early Alzheimer’s disease (AD). Determination of sensitive and specific markers of very early AD progression is intended to aid researchers and clinicians to develop new treatments and monitor their effectiveness, as well as lessen the time and cost of clinical trials.

The Principal Investigator of this initiative is Michael W. Weiner, MD, VA Medical Center, and University of California-San Francisco. ADNI is the result of efforts of many co-investigators from a broad range of academic institutions and private corporations, and subjects have been recruited from over 50 sites across the U.S. and Canada. The initial goal of ADNI was to recruit 800 subjects but ADNI has been followed by ADNI-GO and ADNI-2. To date these three protocols have recruited over 1500 adults, ages 55 to 90, to participate in the research, consisting of cognitively normal older individuals, people with early or late MCI, and people with early AD. The follow up duration of each group is specified in the protocols for ADNI-1, ADNI-2 and ADNI-GO. Subjects originally recruited for ADNI-1 and ADNI-GO had the option to be followed in ADNI-2.

**IMAS Methods**

The Indiana Memory and Aging Study (IMAS) is a longitudinal observational study of older adults at risk for and with clinical Alzheimer’s disease. As part of the IMAS study, participants underwent a comprehensive phone screen for comorbid medical conditions and a comprehensive clinical visit through the Indiana Alzheimer Disease Research Center (IADRC), which included medical history, clinical and neurologic testing, neuropsychological testing with the Uniform Dataset 3 (UDS-3) battery [1], a blood sample, and a structural and functional MRI. Diagnoses were made using similar criteria as used in ADNI. Specifically, patients were diagnosed with AD and MCI using standard criteria [2, 3], while CN participants had no significant memory concerns or performance deficits on cognitive testing. In addition, IMAS includes individuals diagnosed as subjective cognitive decline (SCD) according to the following criteria: elevated levels of subjective memory concerns on the 20-item Cognitive Change Index (CCI-20), reflected as a score of 20 or more on the first 12 items, with or without increased levels of informant-based concerns [4, 5], and without a measurable cognitive deficit.

**Supplemental Tables**

**Supplemental Table S1**. ADNI Demographics for Field Strength Analyses.

|  | 1.5T Scan 1 (n=75) | 1.5T Scan 2 (n=73) | 3T Scan 1 (n=75) | 3T Scan 2 (n=74) |
| --- | --- | --- | --- | --- |
| Diagnostic Group (CN/MCI/AD) | 25/25/25 | 25/24/24 | 25/25/25 | 24/25/25 |
| Sex (M/F) | 31/44 | 30/43 | 31/44 | 30/44 |
| Age (years) | 74.47 (6.66) | 74.53 (6.69) | 74.47 (6.66) | 74.31 (6.57) |
| Age range (low-high) | 59-89 | 59-89 | 59-89 | 59-89 |
| Education (years) | 15.80 (2.89) | 15.71 (2.87) | 15.80 (2.89) | 15.74 (2.86) |
| Handedness (right/left) | 69/6 | 67/6 | 69/6 | 68/6 |
| Brain Age | 74.62 (9.69) | 74.74 (9.94) | 70.49 (9.87) | 70.02 (9.45) |
| BAG | 0.16 (8.55) | 0.21 (8.57) | -3.98 (8.70) | -4.29 (8.42) |
| Brain Age after ComBat | 72.53 (9.74) | 72.66 (10.01) | 72.57 (9.93) | 72.09 (9.52) |
| Brain Age after transformation | 74.62 (9.69) | 74.74 (9.94) | 74.57 (10.39) | 74.07 (9.94) |

ADNI participants (25 CN / 25 MCI / 25AD) underwent two MRI scans in a 1.5T field strength scanner and two scans in a 3T scanner. While these are the same individuals repeated in each scan, due to missed scans or failed scans that were excluded, there are small differences in the number of participants in between each group. Group demographics, brain age estimates before and after ComBat and the linear transformation, and brain age gap (BAG) results by scanner strength and scan number are shown as mean (standard deviation). AD = Alzheimer’s disease; BAG = brain age gap; CN = cognitively normal; F = female; M = male; MCI = mild cognitive impairment

**Supplemental Table S2.** IMAS Demographics for Voxel Size Analyses.

|  | 0.8mm^3^ Scan (n=47) | 1mm^3^ Scan (n=16) | ADNI2 Scan (n=47) |
| --- | --- | --- | --- |
| Diagnostic Group (CN/SCD/MCI/AD) | 19/15/10/3 | 8/5/3/0 | 18/16/10/3 |
| Sex (M/F) | 22/25 | 7/9 | 22/25 |
| Age (years) | 74.21 (7.10) | 75.25 (6.50) | 74.20 (7.09) |
| Age range (low-high) | 61.69-91.92 | 62.78-89.72 | 61.69-91.92 |
| Education (years) | 17.53 (1.94) | 17.81 (1.72) | 17.53 (1.94) |
| Handedness (right/left) | 40/5/1/1 | 14/2/0/0 | 40/5/1/1 |
| Brain Age | 70.69 (11.05) | 71.64 (8.99) | 71.13 (10.45) |
| BAG | -3.52 (8.13) | -3.61 (8.04) | -3.07 (7.63) |

IMAS participants underwent one MRI scan using a 0.8mm^3^ voxel size, one scan using a 1mm^3^ voxel size, and one scan using the ADNI2 sequence (1.1 x 1.1 x 1.2 mm) on a Siemens Prisma 3T scanner. Demographics, brain age estimations, and brain age gap (BAG) results are shown as mean (standard deviation). AD = Alzheimer’s disease; BAG = brain age gap; CN = cognitively normal; F = female; M = male; MCI = mild cognitive impairment; SCD = subjective cognitive decline

**Supplemental Table S3.** IMAS Demographics: For Race/Ethnicity Analyses.

|  | NHW (n=71) | AA (n=71) | Group Difference |
| --- | --- | --- | --- |
| Diagnostic Group (CN/SCD/MCI/AD/AD-vascular) | 30/19/15/6/1 | 30/19/15/7/0 | ꭓ = 1.077 (p=0.898) |
| Sex (M/F) | 17/54 | 17/54 | ꭓ = 0 (p=1) |
| Age (years) | 68.87 (8.39) | 69.07 (8.41) | t= -0.140 (p=0.889) |
| Age range (low-high) | 47-91 | 52-90 | - |
| Education (years) | 15.55 (2.31) | 14.90 (2.51) | t = 1.599 (p=0.112) |
| Handedness (right/left/ambidextrous) | 68/2/1 | 66/5/0 | ꭓ = 2.316 (p=0.314) |
| Brain Age | 65.01 (9.79) | 62.58 (10.78) | t = 1.405 (p=0.162) |
| BAG | -3.87 (6.79) | -6.49 (8.50) | t = 2.034 (p=0.044) |

IMAS participants (71 non-Hispanic White (NHW), 71 African American (AA)) were matched as closely as possible on diagnostic group, sex, age, education, and handedness. There were no differences in any demographic variables between NHWs and AAs. Raw brain age estimation did not differ between the groups; however, brain age gap (BAG) was significantly different between the NHWs and AAs. AA = African American; AD = Alzheimer’s disease; AD-vascular = Alzheimer’s disease with vascular contribution; BAG = brain age gap; CN = cognitively normal; F = female; M = male; MCI = mild cognitive impairment; NHW = non-Hispanic White; SCD = subjective cognitive decline

**Supplemental Table S4.** ADNI Demographics: For Race/Ethnicity Analyses.

|  | NHW (n=147) | AA (n=147) | Group Difference |
| --- | --- | --- | --- |
| Diagnostic Group  (CN/SMC/EMCI/LMCI/AD) | 41/44/20/27/15 | 41/44/20/27/15 | ꭓ = 0 (p=1) |
| Sex (M/F) | 44/103 | 44/103 | ꭓ = 0 (p=1) |
| Age (years) | 69.65 (7.03) | 69.52 (7.39) | t = 0.162 (p=0.872) |
| Age range (low-high) | 55-88 | 56-89 | - |
| Education (years) | 15.63 (2.23) | 15.57 (2.42) | t = 0.225 (p=0.822) |
| Handedness (right/left) | 138/9 | 134/13 | ꭓ = 0.786 (p=0.375) |
| Brain Age | 65.78 (11.18) | 62.73 (10.24) | t = 2.439 (p=0.015) |
| BAG | -3.87 (8.84) | -6.79 (8.01) | t = 2.963 (p=0.003) |

ADNI participants (147 Non-Hispanic White (NHW), 147 African American (AA)) were matched as closely as possible on diagnostic group, sex, age, education, and handedness. There were no differences in any of the diagnostic variables between NHWs and AAs. Raw brain age estimation and BAG were significantly different between the NHWs and AAs. AA = African American; AD = Alzheimer’s disease; BAG = brain age gap; CN = cognitively normal; EMCI = early mild cognitive impairment; F = female; LMCI = late mild cognitive impairment; M = male; NHW = non-Hispanic White; SMC = significant memory concern

**Supplemental Table S5.** ADNI Demographics: For Race/Ethnicity Analyses.

|  | NHW (n=76) | Hispanic (n=76) | Group Difference |
| --- | --- | --- | --- |
| Diagnostic Group (CN/EMCI/LMCI/AD/SMC) | 24/16/11/11/14 | 24/16/11/11/14 | ꭓ = 0 (p=1) |
| Sex (M/F) | 32/44 | 32/44 | ꭓ = 0 (p=1) |
| Age (years) | 70.08 (7.48) | 69.88 (7.72) | t = 0.160 (p=0.873) |
| Age range (low-high) | 55-90 | 55-90 | - |
| Education (years) | 16.05 (2.27) | 16.17 (2.56) | t = -0.302 (p=0.763) |
| Handedness (right/left) | 75/1 | 74/2 | ꭓ = 0.340 (p=0.560) |
| Brain Age | 67.32 (11.53) | 66.14 (9.90) | t = 0.678 (p=0.499) |
| BAG | -2.76 (8.04) | -3.75 (7.64) | t = 0.773 (p=0.441) |

ADNI participants (76 non-Hispanic White (NHW), 76 Hispanics White) were matched as closely as possible on diagnostic group, sex, age, education, and handedness. There were no differences in any of the diagnostic variables between NHWs and Hispanic Whites. There were also no differences in raw brain age estimation or BAG between the NHWs and Hispanic Whites. AD = Alzheimer’s disease; BAG = brain age gap; CN = cognitively normal; EMCI = early mild cognitive impairment; F = female; LMCI = late mild cognitive impairment; M = male; NHW = non-Hispanic White; SMC = significant memory concern

**Supplemental Table S6.** ADNI Demographics: For Race/Ethnicity Analyses.

|  | NHW (n=47) | Asian (n=47) | Group Difference |
| --- | --- | --- | --- |
| Diagnostic Group (CN/EMCI/LMCI/AD/SMC) | 12/7/4/9/15 | 12/7/4/9/15 | ꭓ = 0 (p=1) |
| Sex (M/F) | 23/24 | 23/24 | ꭓ = 0 (p=1) |
| Age (years) | 71.02 (8.71) | 70.64 (9.38) | t = 0.205 (p=0.838) |
| Age range (low-high) | 59-88 | 57-88 | - |
| Education (years) | 16.85 (2.02) | 16.96 (2.16) | t = -0.247 (p=0.806) |
| Handedness (right/left) | 45/2 | 46/1 | ꭓ = 0.344 (p=0.557) |
| Brain Age | 69.46 (12.25) | 65.05 (10.80) | t = 1.850 (p=0.068) |
| BAG | -1.56 (9.64) | -5.59 (7.80) | t = 2.224 (p=0.029) |

ADNI participants (47 non-Hispanic White (NHW), 47 Asians) were matched as closely as possible on diagnostic group, sex, age, education, and handedness. There were no differences in any of the diagnostic variables between NHWs and Asians. A significant difference in BAG between NHWs and Asians was observed, while only a trend for a statistically significant difference in raw brain age estimation was seen. AD = Alzheimer’s disease; BAG = brain age gap; CN = cognitively normal; EMCI = early mild cognitive impairment; F = female; LMCI = late mild cognitive impairment; M = male; NHW = non-Hispanic White; SMC = significant memory concern
